# MUSCLE SYNERGIES ANALYSIS SHOWS ALTERED NEURAL COMMANDS IN WOMEN WITH PATELLOFEMORAL PAIN DURING WALKING

**DOI:** 10.1101/2022.11.07.22282031

**Authors:** Cintia Lopes Ferreira, Filipe O. Barroso, Diego Torricelli, José L. Pons, Fabiano Politti, Paulo Roberto Garcia Lucareli

## Abstract

**Backgroud:** Several studies suggest that the central nervous system coordinates muscle activation by modulating neural commands directed to groups of muscles combined to form muscle synergies. Individuals with patellofemoral pain (PFP) move differently from asymptomatic individuals. Understanding the neural factors involved in the execution of tasks such as walking can help comprehend how the movement is planned and better understand this clinical condition.

**Research question:** To compare the muscle coordination of women with and without PFP during gait.

**Methods:** Eleven women with PFP and thirteen asymptomatic women were assessed using three-dimensional kinematics and electromyography (EMG) while walking at self-selected speed. Kinematics of the trunk, pelvis and lower limbs were analyzed through the Movement Deviation Profile. Muscle synergies were extracted from the EMG signals of eight lower limb muscles collected throughout the whole gait cycle.

**Results:** Kinematic differences between the two groups (p<0.001, z-score=3.06) were more evident during loading response, terminal stance, and pre-swing. PFP group presented a lower number of muscle synergies (p=0.037), and greater variability accounted for (VAF_total_) when using 3 (p=0.017), 4 (p=0.004), and 5 (p=0.012) synergies to reconstruct all EMG signals. The PFP group also presented higher VAF_muscle_ for rectus femoris (p=0.048) and gastrocnemius medialis (p=0.019) when considering 4 synergies.

**Significance:** Our results suggest that women with PFP show lower motor complexity and deficit in muscle coordination to execute gait, indicating that gait in PFP gait is the result of different neural commands compared to asymptomatic women.

## Introduction

Patellofemoral pain (PFP) is a chronic pain condition characterized by a multifactorial origin [1]. Differences in kinematics [2], kinetics [3] and reduction in neuromuscular efficiency of the quadriceps muscles [4] are observed in individuals with PFP during gait.

Neural control of human locomotion results from the complex interaction between central and peripheral inputs that coordinates the various degrees of freedom of the musculoskeletal system [5]. As a result, similar movements can be produced by different muscle activation and coordination patterns [6]. The hypothesis of muscle synergies suggests that the central nervous system (CNS) does not activate muscles individually but rather sends neural commands that activate specific muscle groups to produce the movement [7]. Thus, the analysis of muscle synergies has been used to unravel neural strategies underlying human locomotion [6].The literature is inconclusive regarding the behavior of muscle synergies in musculoskeletal conditions [8]. Previous studies have shown reduced muscle coordination for muscles close to the painful region in women with PFP during lateral step down (LSD) [9], and in individuals with femoroacetabular impingement [10] and gluteal tendinopathy [11] during gait. By identifying a painful episode as a threat, the CNS seems to modify the motor behavior to control pain and protect the painful region [12]. However, if pain persists in addition to the cognitive-behavioral factors, this alteration in motor behavior can contribute to the persistence and chronification of the pain [13].

Several studies have investigated how the CNS controls pathological and normal human locomotion [14–16]. However, we do not know to date how PFP can interfere in motor control during gait. Therefore, comprehending the possible neural factors underlying gait execution in individuals with PFP can help understand this clinical condition. Based on the literature, we hypothesized that women with PFP exhibit alterations in muscle coordination, mainly in quadriceps muscles, during gait. Therefore, the objective of this study was to compare muscle synergies between women with and without PFP during walking.

## Methods

### Study design

A cross-sectional study was designed, and approved by the research ethics committee of the Nove de Julho University, according to the National Research Ethics Commission of Brazil, in compliance with all applicable Federal regulations governing the protection of human subjects. Research data were derived from an approved by the research ethics committee of the Nove de Julho University, number 2.732.0. and conducted at the Núcleo de Apoio à Pesquisa em Análise do Movimento of the Nove de Julho University. All participants agreed to participate and signed the corresponding informed consent.

### Participants

Eleven women with PFP and thirteen asymptomatic women were selected. The assessment stages of this study were similar to our previous study [9].

Inclusion criteria were: age between 18-35 years and body mass index below 30kg/m^2^. For the PFP group (PFPG), subjects should have experienced PFP for at least three months with a minimum Numerical Pain Rating Scale (NPRS) [17] score of three points at least during two of the following activities: squat, run, ascent and descent stairs, kneeling or sitting for prolonged hours with flexed knees.

Exclusion criteria for both groups were: presence of lateral and/or posterior knee pain, history of ligament and/or meniscal injuries, surgical procedures in the lower limbs or spine, ankle sprain, low back pain, more than one episode of patellar dislocation, cardiorespiratory, neurological or musculoskeletal problems that could interfere or prevent the subject from performing the entire assessment, and taking controlled medication such as antidepressants. The control group (CG) could not present any lower limb musculoskeletal pain.

### Procedures and instrumentation

Assessments were performed on two non-consecutive days, with a maximum interval of 15 days between them.

On the first day, the candidates for the PFPG were evaluated by two physiotherapists to clinically confirm PFP [18]. All selected subjects underwent 3D kinematics and EMG assessment during gait. Kinematic analysis was performed using an eight-camera Vicon system operating at 240 Hz. According to the Plug-in Gait model, twenty-five retroreflective markers were attached to each subject’s skin. EMG signals were captured using an eight-channel wireless acquisition system (EMG System do Brasil Ltda^®^) composed of bipolar active electrodes with a 1k amplification gain and 20-500 Hz bandpass analog filter. EMG signals were recorded with a sampling frequency of 2,400 Hz. Following SENIAM recommendations [19], Ag/AglCl (Miotec^®^) surface electrodes were positioned on the skin, taking into account the following locations: Adductor Longus (AdLo) [20], Gluteus Medius (GlMe), Vastus Lateralis (VaLa), Rectus Femoris (ReFe), Vastus Medialis (VaMe), Biceps Femoris (BiFe), Tibialis Anterior (TiAn) and Gastrocnemius Medialis (GaMe). The limb with pain or the most painful one for the PFPG and the dominant one for the CG was analyzed. All participants walked at self-selected speed on a 6-meter-long by 1-meter-wide track. Data from at least 25-30 strides were collected, where corresponding EMG signals presented no artifacts or noise that would prevent a trustful analysis.

On the second day, the isometric strength test was performed on hip abductors and lateral rotators, as well as on hip and knee extensors, using a manual dynamometer (Lafayette, IN, USA) [21]. Foot posture index [22] and lunge test [23] were also performed. All subjects answered the questionnaires on the quality of life (SF-36-Medical Outcomes Study 36 – Item Short-Form Health Survey) [24], depression (BECK Depression Inventory) [25], and function (Anterior Knee Pain Scale – AKPS) [17]. The PFPG also answered questionnaires related to the intensity of pain in the last 15 days and during gait assessment (NPRS) [17], symptom duration, catastrophizing (Pain Catastrophizing Scale) [26], and kinesiophobia (Tampa Scale of Kinesiophobia) [27].

### Data Analysis

Two researchers with experience in EMG analysis and with no involvement in the data collection process guaranteed, by visual inspection, the quality of EMG data of the eight muscles for 25-30 strides per subject.

The 3D marker trajectories were processed using the Vicon Nexus 2.10 software to estimate the joint centers [28]. The strides were identified according to Lopes Ferreira et al., 2019 [2]. A Woltring filter with a 2 mean square error was applied to reduce the vibratory noise during the marker trajectories due to soft tissue artifacts.

The absolute angles from frontal, sagittal, and transverse planes of the trunk and pelvis segments in relation to the laboratory; frontal, sagittal, and transverse plane of the hip in relation to the pelvis; frontal and sagittal planes of the knee to the thigh; sagittal plane from the foot to the shank; and absolute transverse plane of the foot were analyzed through the Movement Deviation Profile (MDP) [29]:. An MDP curve consisting of normalized data points every 2% of the movement cycle was calculated using the CG data. The average of the 51 points of the MDP curve of all strides was considered for statistical comparison between groups for each gait cycle. The linear parameters: stride and step time, normalized cadence, percentage of first and second double support, single support, stance phase, normalized step length, and speed of each stride were analyzed.

Raw EMG data were exported from the Vicon Nexus software and processed with MATLAB R2018a (The MathWorks, Natick, MA, USA). Concatenated raw EMG signals from each participant were high-pass filtered at 20 Hz, rectified and low-pass filtered at 5 Hz to obtain the EMG envelopes [30]. For each muscle and subject, EMG envelopes were amplitude-normalized by the average of the peaks of each stride and time-normalized by resampling EMG envelopes at each 1% of the cycle [31]. For each subject, normalized EMG envelopes were combined into *m* x *t* (EMG_o_) matrices, where *m* is the number of muscles (eight in this study) and *t* is the time base [number of gait cycles (25-30) x 100] [16]. Muscle synergies components were calculated applying a non-negative matrix factorization (NNMF) algorithm [32]. Mathematically, the algorithm is described by the following equation:

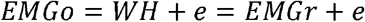

Where *W* is the *m* x *n* matrix specifying the weight of each muscle in each synergy, *n* is the number of synergies, *H* is the *n* x *t* matrix specifying the time-varying activation coefficients representing the recruitment of each synergy throughout the cycle. EMG_r_ is the matrix *m* x *t* resulting from the multiplication of *W* and *H* (reconstructed EMG envelopes), and *e* is the residual error [33]. We considered 3 to 5 synergies (*n* = 3, 4, 5) as input to the NNMF algorithm. For each *n*, NNMF was run 40 times, and the repetition with the smallest reconstruction error was selected [16].

The minimum number of synergies required to guarantee adequate reconstruction of the EMG signal was determined as the minimum number necessary to obtain variability account for (VAF_total_) ≥90%, as well as VAF_muscle_ ≥75% for all muscles assessed [30].

CG data were used to obtain two reference matrices: *W*_*ref*_ (representing the contribution of each muscle to each synergy) and *H*_*ref*_ (containing the activation coefficients and representing how each synergy is modulated over time). For this purpose, EMG_o_ matrices of all asymptomatic subjects were concatenated, and the NNMF algorithm was applied to obtain *W*_*ref*_ and *H*_*ref*_ for 3, 4, and 5 synergies. Then, for each PFP subject, synergy vectors (columns of matrices *W*) were ordered based on their similarity with synergy vectors from *W*_*ref*_ using normalized scalar product. After ordering the synergy vectors (*W*) and the corresponding activation coefficients (*H*), the grand average of *W* and *H* for each group was calculated to compare both groups.

### Statistical Analysis

The Shapiro-Wilk test was used to verify the distribution of the analyzed data. When data were considered parametric, the t-test for independent samples was performed, and when non-parametric, the Mann-Whitney test was used to compare the groups. Data were expressed as a mean and confidence interval (CI 95%). A statistically significant *p*-value was considered when *p*≤0.05. Z-score was calculated to quantify the difference in MDP between the groups [2].

The statistical parametric mapping (SPM) two-tailed independent t-test was used to compared EMG envelopes of each muscle during all normalized gait cycles between groups. SPM uses a random field theory correction to ensure that only values ≥ 5% of the data points of the SPM {t} reach the significance threshold (α = 0.05) by chance if the trajectory of the SPM {t} results from an equally smooth random process. Finally, the probability (p) value was calculated for each supra-threshold and infra-threshold region when SPM {t} exceeded the critical threshold [34]. All SPM analyses were performed with an spm1d open code (www.spm1d.org).

## Results

The sample characterization data and the spatiotemporal gait parameters are shown in Table 1 and 2, respectively. PFPG showed worse scores of AKPS and physical functioning, role physical, bodily pain, and vitality domains of the SF-36 questionnaire and walked with a greater step length and cadence.

**Table 1.** Mean and confidence interval (95%) of demographic data, physical assessments and questionnaires scores, and comparison between the groups.

|  | Control Group<br>mean (CI 95%) | PFP Group<br>mean (CI 95%) | P |
| --- | --- | --- | --- |
| Age (years) | 24.15 (21.14-27.16) | 24.18 (21.33-27.03) | 0.988 |
| Height (m) | **1.61 (1.58-1.64)** | **1.66 (1.63-1.69)** | **0.011** |
| Body mass (kg)<sup>+</sup> | 56.50 (51.47-61.53) | 64.45 (55.65-73.26) | 0.082 |
| BMI (kg/m<sup>2</sup>) | 21.86 (19.90-23.81) | 23.37 (20.12-26.62) | 0.371 |
| HABD (%BW) | 19.62 (15.30-23.93) | 16.38 (12.20-20.57) | 0.254 |
| HEXT (%BW) | 21.46 (16.00-26.93) | 19.91 (13.47-26.34) | 0.687 |
| HLR (%BW) | 10.16 (8.42-11.90) | 9.94 (8.03-11.84) | 0.850 |
| KEXT (%BW) | 34.80 (28.91-40.69) | 32.86 (26.69-39.04) | 0.623 |
| Lunge test (°)<sup>+</sup> | 41.41 (37.18-45.64) | 41.69 (36.94-46.44) | 0.977 |
| FPI | 5.08 (3.65-6.50) | 4.27 (2.21-6.33) | 0.476 |
| BECK (0-63)<sup>+</sup> | 6.69 (2.45-10.94) | 6.82 (4.90-8.74) | 0.450 |
| AKPS (0-100)<sup>+</sup> | **99.69 (99.24-100.15)** | **75.27 (69.91-80.63)** | **0.000** |
| SF-36 |  |  |  |
| *Physical functioning (0-100)<sup>+</sup>* | **94.23 (88.08-100.38)** | **74.55 (63.36-85.73)** | **0.000** |
| *Role physical (0-100)<sup>+</sup>* | **98.08 (93.89-102.27)** | **77.45 (63.50-91.41)** | **0.001** |
| *Bodily pain (0-100)<sup>+</sup>* | **83.85 (77.75-89.94)** | **66.09 (56.87-75.31)** | **0.002** |
| *General health (0-100)* | 79.38 (68.07-90.70) | 70.27 (58.03-82.51) | 0.242 |
| *Vitality (0-100)* | **61.92 (49.43-74.41)** | **41.00 (29.10-52.90)** | **0.015** |
| *Social functioning (0-100)<sup>+</sup>* | 81.73 (66.12-97.34) | 70.00 (54.42-85.57) | 0.108 |
| *Role emotional (0-100)<sup>+</sup>* | 74.36 (50.88-97.84) | 60.00 (33.88-86.11) | 0.276 |
| *Mental health (0-100)* | 73.23 (63.15-83.31) | 60.00 (49.66-70.34) | 0.057 |
| PCS (0-52) | - | 11.18 (7.27-15.10) | - |
| TAMPA (17-68) | - | 30.64 (26.87-34.40) | - |
| NPRS last 15 days (0-10) | - | 5.18 (4.34-6.02) | - |
| NPRS during gait (0-10) | - | 1.91 (0.55-3.27) | - |
| Symptom duration (months) | - | 59.09 (20.51-97.67) | - |

Bold indicates statistically relevant difference $p \leq 0.05$ ; +: Non-parametric data, Mann-Whitney test was used; CI: confidence interval; m: meters; kg: kilogram; BMI: body mass index; kg/m<sup>2</sup>: kilograms per meter square; %BW: body weight percentage; HADB: hip abductors; HEXT: hip extensors; HLR: lateral hip rotators; KEXT: knee extensors; FPI: foot posture index; BECK: Beck depression inventory; AKPS: anterior knee pain scale; SF-36: 36-item short-form health survey questionnaire; PCS: pain catastrophizing scale; TAMPA: Tampa scale of kinesiophobia; NPRS: numerical pain rating scale.

**Table 2.**
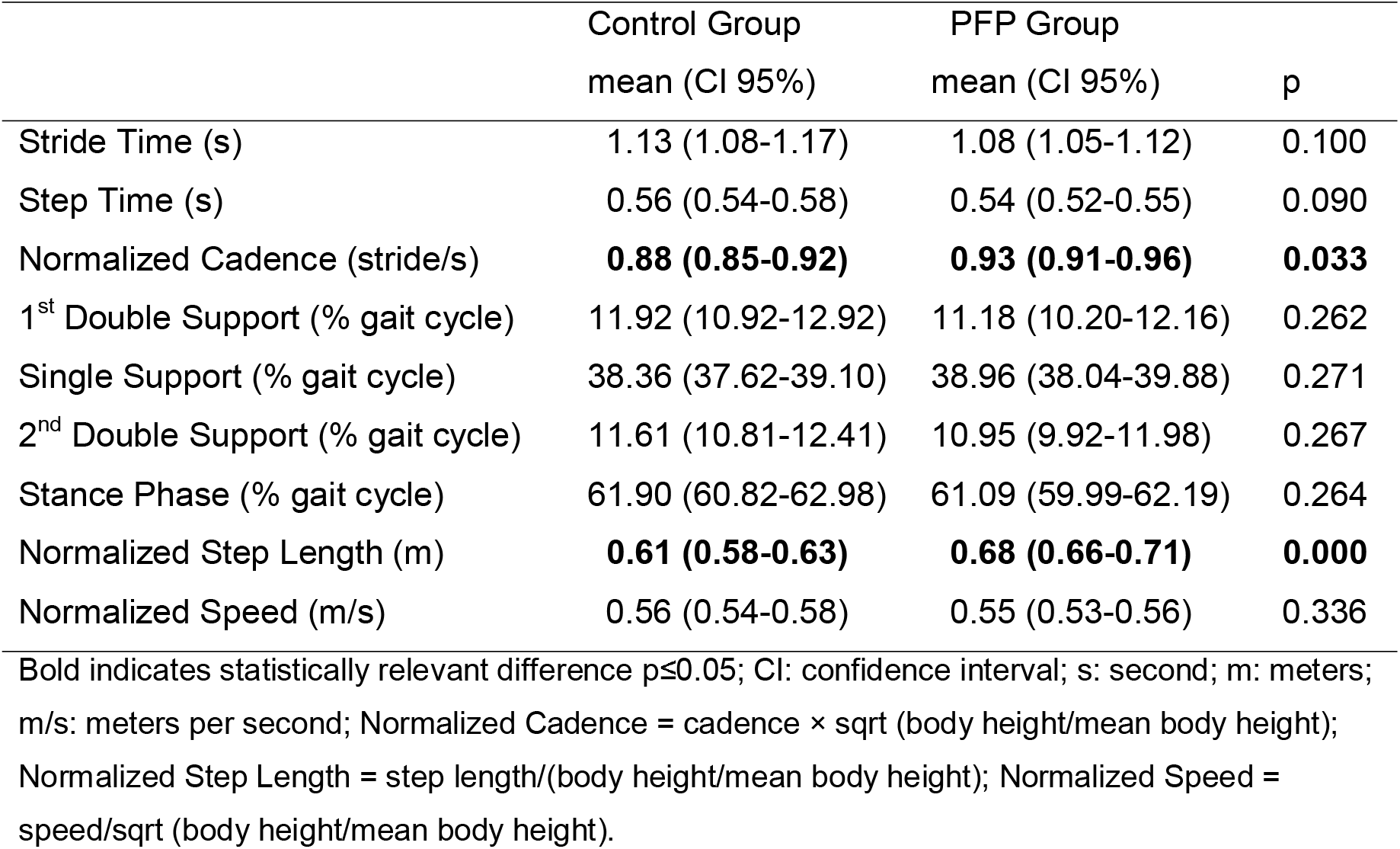
Mean and confidence interval (95%) of spatiotemporal gait parameters and comparison between the groups.

There was a significant difference in the mean (standard deviation) MDP curve between CG 10.4º (1.2º) and PFPG 14.09º (1.4º) (p < 0.001 and z-score = 3.06). MDP analysis showed differences in kinematic behavior between 2% and 12% (loading response), and between 38% and 54% (terminal stance and pre-swing) of the gait cycle, based on 95% confidence interval (Figure 1).

**Figure 1.**
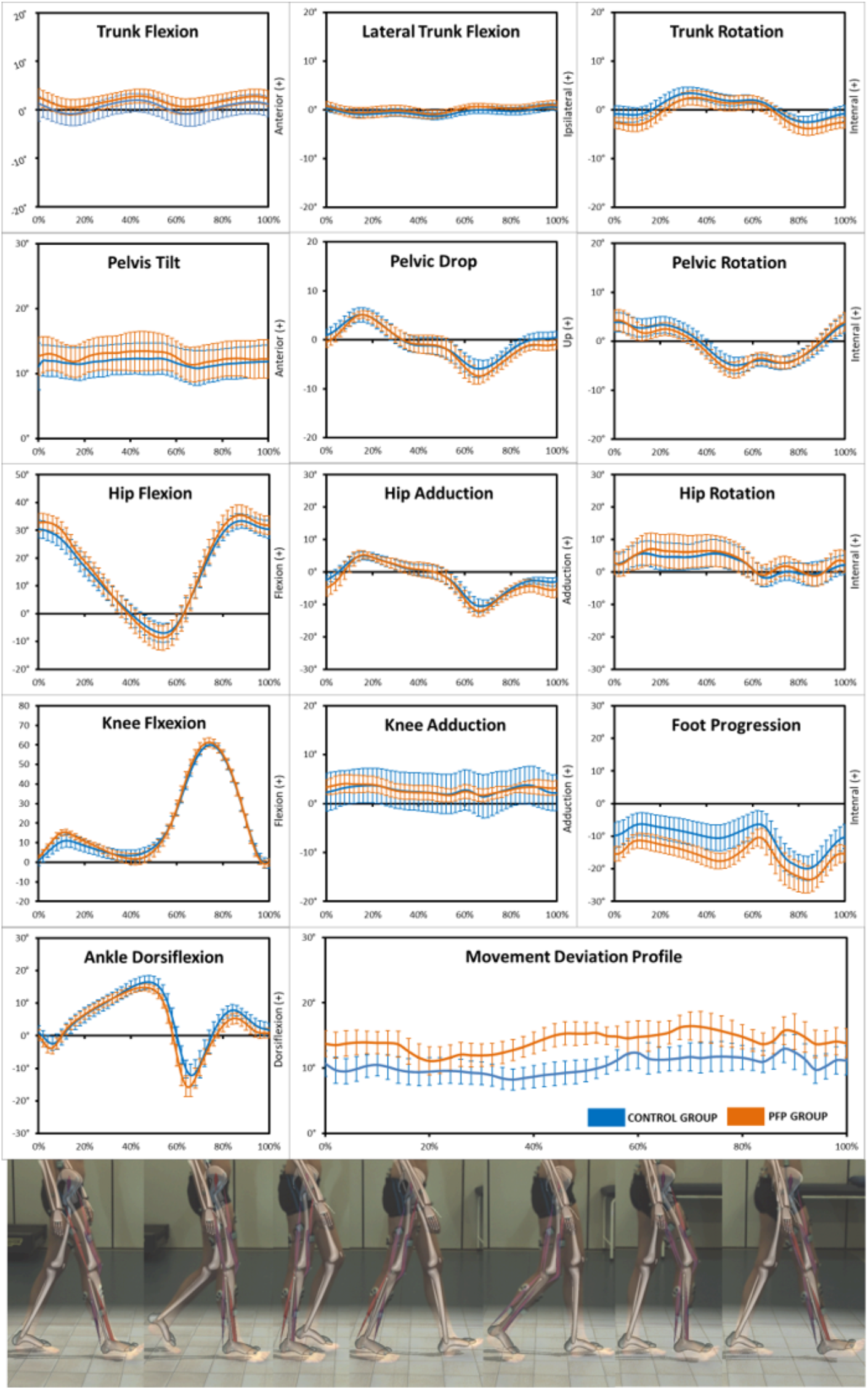
The Kinematics Data from sagittal, frontal, and transverse planes of the trunk, pelvis, hip, knee, and ankle during the entire gait cycle. Movement Deviation Profile (MDP) chart (mean and 95% confidence intervals bands) summarizes the 13 angles curves of kinematics data analyzed for the control group (blue) and PFP group (orange) during gait.

SPM analysis did not find significant differences of individual muscles activation between groups (Figure 2). However, muscle synergies analysis revealed that PFPG presented higher VAF_total_ values when reconstructing EMG signals with 3, 4, and 5 synergies, and higher VAF_muscle_ values for ReFe and GaMe when considering 4 synergies (Table 3).

**Table 3.**
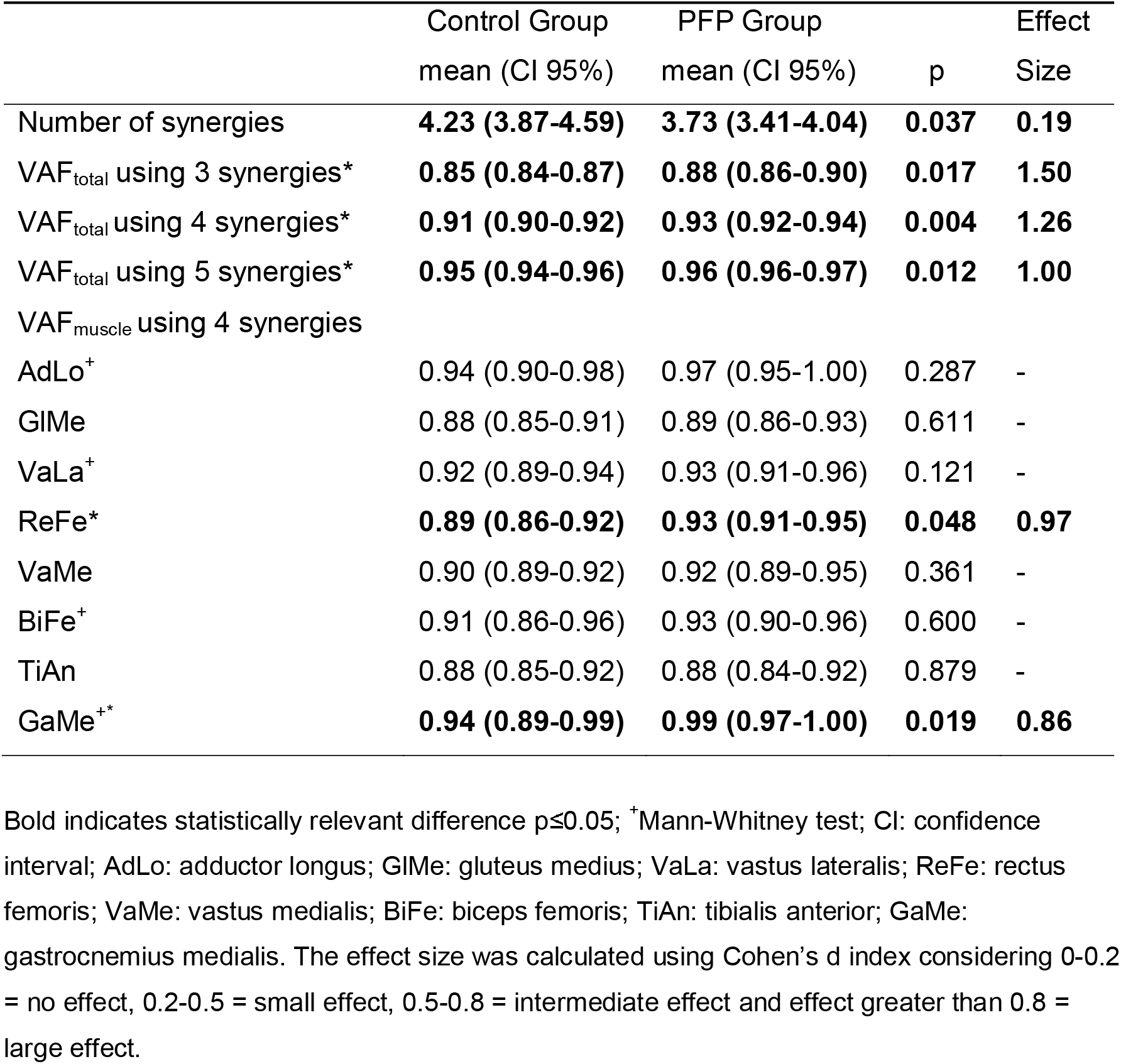
Mean and confidence of interval (95%) of number of synergies, VAF_total_ considering 3, 4, and 5 synergies and VAF_muscle_ for 4 synergies and comparison between groups.

**Figure 2.**
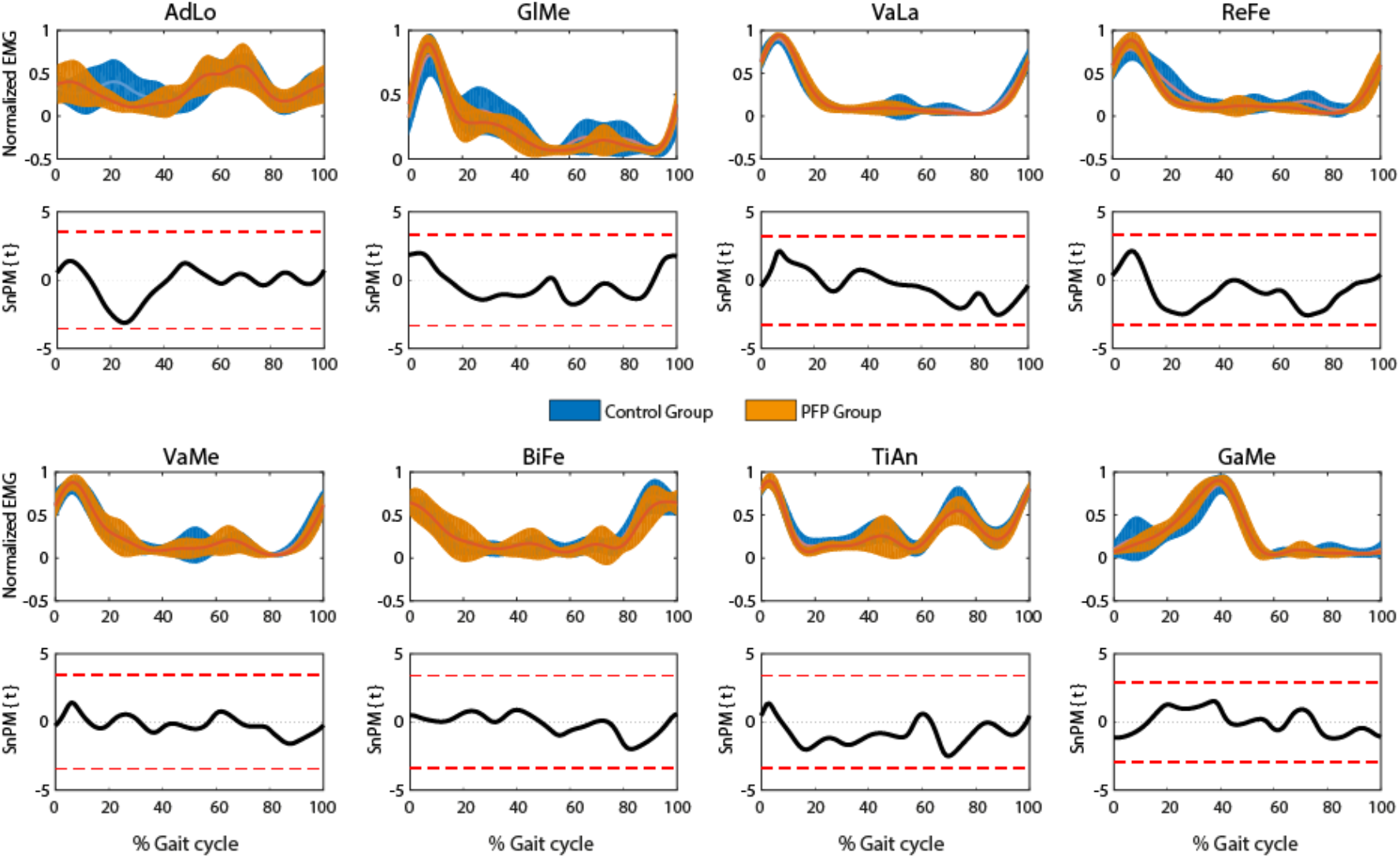
The EMG envelopes from muscle assessed in the first and third row and the SPM analyzes in the second and fourth row during the entire gait cycle for the control group (blue) and PFP group (orange). AdLo: adductor longus; GlMe: gluteus medius; VaLa: vastus lateralis; ReFe: rectus femoris; VaMe: vastus medialis; BiFe: biceps femoris; TiAn: tibialis anterior; GaMe: gastrocnemius medialis.

The minimum number of synergies needed to explain the variability of the EMG signals during gait was different between the groups (Table 3). In CG, 3 synergies were needed to properly reconstruct the EMG signals in one participant, whereas eight participants needed 4 synergies, and three participants needed 5. In PFPG, three patients needed 3 synergies to reconstruct the EMG signals, and eight patients needed 4 synergies.

According to our criteria (VAF_total_ ≥ 90% and VAF_muscle_ ≥ 75%) to determine the minimum number of synergies, we considered 4 synergies to compare the groups (Figure 3). Synergy 1 was active during mid and terminal swing, initial contact, and loading response (H1), and the muscles mainly contributing to this synergy were BiFe and TiAn (W1). Synergy 2, represented by the activity of GlMe, VaLa, ReFe and VaMe (W2), reached its activation peak during the loading response and part of midstance (H2). Synergy 3 had the main contribution of GaMe (W3) and was active during mid and terminal stance, and pre-swing (H3). Initial and mid-swing were controlled by synergy 4 (H4), composed by AdLo and TiAn (W4). There were no differences between groups in terms of recruitment of each synergy throughout the gait cycle (Hs) and the weight of each muscle in each synergy (Ws) (Figure 3).

**Figure 3.**
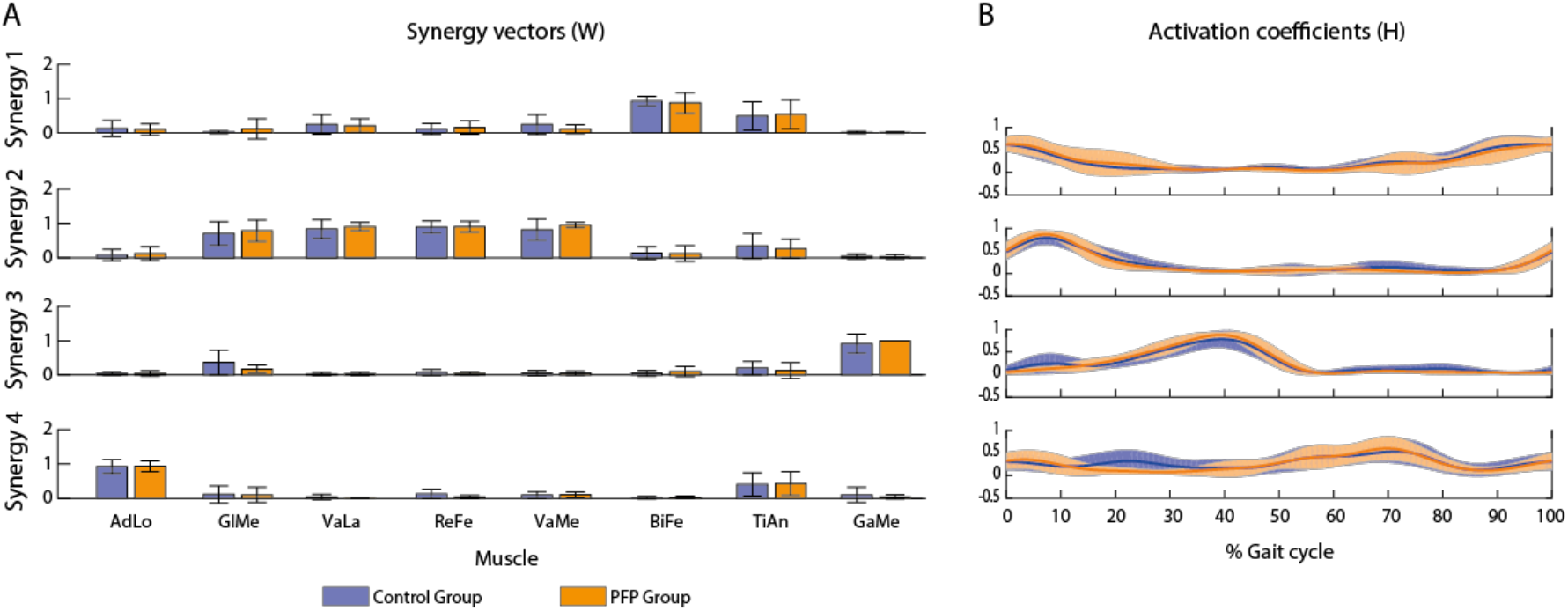
The weight of each muscle in each synergy (Ws) **(A)** and the recruitment of each synergy throughout the gait cycle (Hs) **(B)** during the gait cycle considering 4 synergies. AdLo: adductor longus; GlMe: gluteus medius; VaLa: vastus lateralis; ReFe: rectus femoris; VaMe: vastus medialis; BiFe: biceps femoris; TiAn: tibialis anterior; GaMe: gastrocnemius medialis.

## DISCUSSION

This study compared the coordination of lower limb muscles in women with and without PFP while walking at self-selected speed. Our results add new evidence that women with PFP present reduced motor complexity and muscle coordination during gait (PFPG presented higher VAF_total_ and VAF_muscle_ for ReFe and GaMe) in comparison with women without PFP. This confirms our hypothesis that women with PFP exhibit alterations in muscle coordination.

A lower number of synergies indicates less complexity in terms of muscle coordination and can be associated with increased co-activation levels between muscles and with coupling and fusion mechanisms of synergies in patients with neural injuries [16,35]. In this study, PFPG presented a lower number of synergies during gait. Similar results were observed in individuals with stroke [30,36], spinal cord injury [16] and cerebral palsy [15]. Nevertheless, most of the studies on musculoskeletal disorders do not usually report differences in the number of synergies during gait [8]. The presence of pain on the assessment day and the duration of symptoms reported by the PFPG (59.09 months) can partly justify the results obtained. In painful situations, the CNS seems to redistribute activity intra and between the muscles and generate adaptations in motor variability to protect the musculoskeletal system from movements associated with a painful experience [37].

Different tasks may demand different complexity in motor coordination, which can be assessed by the number of synergies extracted [38]. While we observed differences in the number of synergies between PFPG and CG during gait, the same was not observed in LSD task [9]. We can infer that due to frequent execution of gait as an activity of daily living, the CNS has more opportunities to readjust the musculoskeletal system and make neuromuscular adaptations than in other movements less frequently executed.

It is worth noting that SPM analysis did not find differences in muscle activation between groups for each muscle assessed (Figure 2). SPM compares the mean of the linear envelopes of the EMG signals from each subject of each group. On the contrary, the analysis of synergies reconstructs all EMG signals with basic patterns. The quality of reconstruction is assessed with indicators like VAF_total_ and VAF_muscle_ [6]. Results indicated poorer muscle coordination (higher VAF_total_) in PFPG, which is in line with other findings in tasks like LSD in PFP [9] and gait in patients with femoroacetabular impact [10], gluteal tendinopathy [11] and experimentally induced pain in the low back and calf [39]. The reduced muscle coordination presented by the PFPG mainly occurred in the ReFe and GaMe muscles (higher VAF_muscle_ in PFPG).

ReFe, GlMe, VaLa and VaMe composed synergy 2, which was mainly active in the loading response, and kinematic differences were found at this moment in the gait cycle (Figure 1). Literature has shown that local muscles at regions where the pain is reported present reduced muscle coordination [9–11]. Furthermore, individuals with PFP present local hyperalgesia in the knee, despite not being related to the knee’s muscle strength and angular kinematics [40], which may also contribute to a different neuromuscular behavior on these subjects. Individuals with PFP also present less representation of the quadriceps in the primary motor cortex, indicating impairment in the intermuscular control and coordination and the adoption of more simplified movement strategies for these patients [41]. Finally, it was observed in animal models that the CNS reorganizes muscle activity of the quadriceps to avoid excessive stress and joint tension at the knee joint [42,43].

PFPG also presented poorer muscle coordination of the GaMe (higher VAF_muscle_), which was the main muscle composing synergy 3 and responsible for double support (Figure 3). GaMe is a biarticular muscle that assists in controlling the ankle in the single support and propulsion in second double support [44], and is also a knee flexor. GaMe can be influenced by neuromechanical adaptations that individuals with PFP present after experiencing pain.

Differences in kinematic behavior in the PFPG can be explained by changes in neural control rather than by clinical variables, as both groups had similar muscle strength, dorsiflexion range of motion (lunge test), and foot posture. The CNS seems to adopt different solutions to protect the painful region, and the differences in the VAF_total_ and VAF_muscle_ indicate impairment in the neural control of the movement in painful conditions [39].

The present study has some limitations. The number of assessed muscles does not represent all the muscles activated to execute gait, and this can directly affect the calculation of the number of muscle synergies to describe the task. However, we selected the muscles that exert a primary function in the three main joints of the lower limbs, providing an overall panorama of the task. Our results do not allow us to infer the cause or effect of PFP but provide information on how these patients move and the possible neural factors involved in this condition, which are not possible to identify in the conventional and individual analysis of the EMG signal. Finally, the sample size is extremely small, which again begs the question regarding generalizability. In the long term, prospective studies investigating the influence of neural factors in the appearance and permanence of the PFP and clinical trials approaching these factors in the treatment plan of these patients can better elucidate the importance of the study of these muscle synergies in this group of patients.

## Conclusion

Women with PFP present lower motor complexity and deficit in muscle coordination of the lower limb, indicating that gait in PFP is the result of different neural commands compared to asymptomatic women.

## Data Availability

All data produced in the present study are available upon reasonable request to the authors.

## Acknowledgement

The authors would like to thank the Universidade Nove de Julho (UNINOVE) for providing the evaluation facilities used in the present study.

## Notes

### Competing Interest Statement

The authors have declared no competing interest.

### Funding Statement

The study was financed in part by the Coordenacao de Aperfeicoamento de Pessoal de Nivel Superior; Brasil (CAPES); Financial Code 001.

### Author Declarations

The study was approved by the research ethics committee of the Universidade Nove de Julho, according to the National Research Ethics Commission of Brazil, in compliance with all applicable Federal regulations governing the protection of human subjects. Research data were derived from an approved by the research ethics committee of the Nove de Julho University, protocol, number 2.732.0. All necessary patient/participant consent has been obtained and the appropriate institutional forms have been archived.

